# Identification of the putative causal risk factors and biomarkers of stroke using large-scale genome-wide studies

**DOI:** 10.1101/2023.03.08.23287006

**Authors:** Tania Islam, Luis M García-Marín, Miguel E. Rentería, Gabriel Cuellar-Partida, Asaduzzaman Khan, Jian Zeng, Mohammad Ali Moni

**Affiliations:** School of Health and Rehabilitation Sciences, The University of Queensland, QLD, Australia; Brain and Mental Health Program, QIMR Berghofer, QLD, Australia; School of Biomedical Sciences, Faculty of Medicine, The University of Queensland, Brisbane, QLD, Australia; Translational Research Institute, The University of Queensland, QLD, Australia; Institute for Molecular Bioscience, The University of Queensland, QLD, Australia

**Keywords:** Stroke, latent causal variable method, colocalization, and genome-wide association studies (GWAS)

## Abstract

Stroke is a complex neurological disorder, and the risk factors and genetic biomarkers associated with its development are not completely understood. This study aims to identify putative causal traits and their biomarkers that influence the risk of stroke. We leveraged genome-wide association studies (GWAS) datasets to understand potential causal genetic relationships between stroke and 1,504 complex traits via the latent causal variable (LCV) and Generalised Mendelian randomisation (GSMR) methods. Our results suggest 14 traits showing potential causal genetic effects with stroke risk (|GCP|> 0.60; FDR < 0.05). These traits include cardiovascular, metabolic, and blood clot-related traits. Using mBAT-combo, we identified genes associated with these putatively causal traits and stroke, suggesting shared genetic architectures. Colocalisation analysis showed that several of those overlapping genes were colocalised between stroke and risk traits. Functional enrichment analyses of the overlapping genes highlight the role of coagulation and complement systems, specifically prothrombin and platelet activation, as well as complement and coagulation cascades. This study suggests putative causal genetic architecture and biological pathways shared between stroke and its causal traits.

## Introduction

Stroke is the second largest cause of mortality and disability worldwide [1]. Stroke affects around 13.7 million individuals, of which 5.5 million die yearly [2]. Stroke is a neurological condition characterized by the blockage of blood vessels in the brain, obstructing blood flow and rupture of blood vessels, resulting in hemorrhage in the brain. When the brain arteries rupture during a stroke event, neural demise is observed as a result of the lack of available oxygen [2].

Stroke is a heritable trait, as evidenced by past twin studies and family history [3]. The heritability for ischemic stroke has been estimated at 37.9% [4]. In addition, genome-wide association studies (GWAS) have identified >80 independent risk loci that have been associated with stroke in European and cross-ancestry populations [5]. However, the causal genetic architecture of stroke remains not understood due to high linkage disequilibrium (LD) structures and the influence of numerous environmental factors, including hypertension, metabolic and cardiovascular diseases, associated with increased risk of stroke, which are evident from various observational studies [6, 7]. These findings are broadly compatible with genetic studies, which support the causal genetic role of multiple risk factors in stroke pathophysiology. However, genetic evidence also highlights additional complexities, including the interplay of genetic predispositions and environmental factors, further underscoring the multifactorial nature of stroke risk. Mendelian randomization (MR) and latent causal variable (LCV) are methods frequently used in genetic epidemiology to assess potential causal genetic effects among genetically correlated phenotypes using GWAS data [8, 9]. The identification of putative causal effects between traits is commonly interpreted based on vertical and horizontal pleiotropy [10]. When genetic variants directly influence trait A and trait B, this type of effect is called horizontal pleiotropy. On the other hand, vertical pleiotropy may be seen as a causal cascade where the influence of a genetic variant on a single trait is explained by its impact on another trait [8]. Horizontal pleiotropy could lead to high-false positive results in genetic epidemiology to assess the causality [11, 12].

The LCV method offers several advantages compared to traditional MR methods, though it is important to recognise that no single approach is without limitations. Unlike the conventional MR methods, LCV provides a framework for distinguishing between genetic correlation and potential genetic causality, offering insights into whether observed genetic overlap reflects direct causal relationships or shared genetic architecture [13]. Additionally, LCV estimates the genetic causality proportion (gcp-value), quantifying the degree of causation in a way that is less likely to be confounded by genetic correlations than methods like MR Egger [11]. Recent advancements, such as the generalized summary data-based Mendelian randomization (GSMR) approach, also address some limitations of traditional MR methods by leveraging more SNPs as instrumental variables and employing the HEIDI outlier method to minimize the impact of pleiotropy[13].

Previous studies have significantly advanced our understanding of stroke, identifying various genetic, lifestyle, and clinical factors that contribute to its risk. However, despite these advancements, several gaps remain largely unrecognised. The causal relationships between many identified risk factors and stroke are still not fully understood, particularly the extent to which shared genetic architecture versus direct causal pathways contribute to stroke risk. Therefore, there is a growing interest in exploring the potential causal genetic risk factors of stroke. In the present study, we performed exploratory large-scale genetic screening for potential risk factors of stroke and shared gene biomarkers and underlying pathways for the causal traits and stroke.

## Methods

The overall analytical approach that we employed in this study is shown in Figure 1.

**Figure 1.**
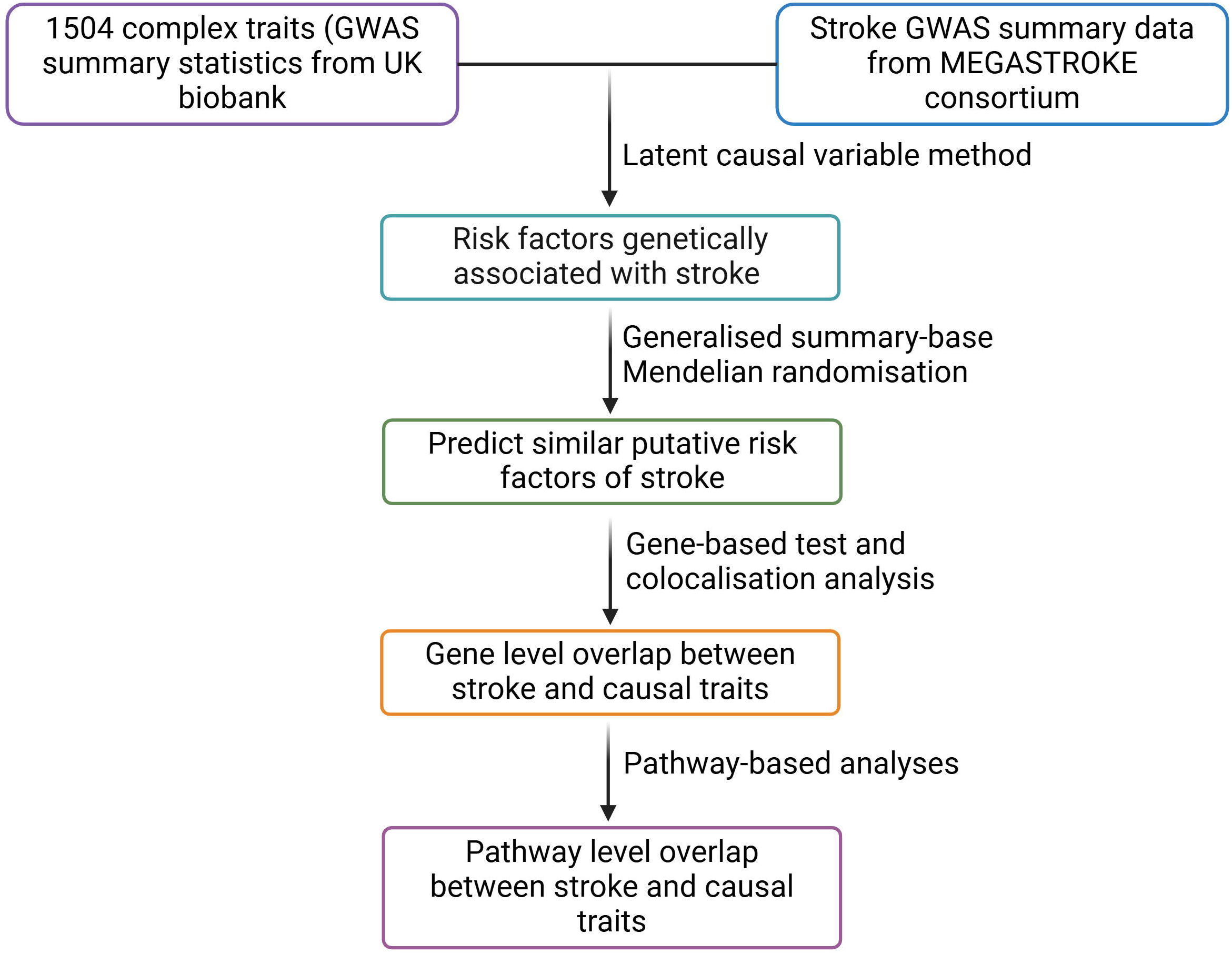
This schematic diagram illustrates the overall study design. Linkage disequilibrium score (LDSC) regression and latent causal variable (LCV) analyses were performed to calculate genetic correlations and causal association between stroke and 1,504 complex traits. The generalized Mendelian randomization (GSMR) method was applied to the result of the LCV method which also revealed that 14 traits were causally associated with stroke. Gene-based tests, colocalization, and pathway-based analyses identified shared genetic architecture and molecular mechanisms between these causal traits and stroke, offering insights into potential pathways driving stroke risk.

### Stroke GWAS data

We obtained GWAS summary statistics for stroke from a meta-analysis of GWAS data from MEGASTROKE consortium (http://megastroke.org/). This data was generated based on the fixed-effects meta-analysis including 40,585 stroke cases and 406,111 controls of European ancestry. We performed the rigorous quality control of the GWAS data and exclude the SNPs with a minor allele frequency MAF < 1% to ensure the robustness of the data. The details of the study have been described in [14].

### UK biobank cohort datasets

We used 1504 phenotypes available at the Complex Traits Genetics Virtual Lab (CTG-VL) web portal (https://genoma.io) [15], which predominantly includes GWAS from the UK Biobank generated by the Neale Lab (http://www.nealelab.is/uk-biobank). Thus, most of these GWAS summary results represent people of European ancestry. These data were corrected for age, age-squared, inferred sex, and genetic ancestry [15].

### Genetic correlations and latent causal variable

The latent causal variable (LCV) method primarily estimates genetic correlation between two traits by applying a linkage disequilibrium score regression (LDSC) method. LDSC is a robust statistical method that leverages genome-wide summary statistics to estimate SNP-based heritability and evaluate the genetic correlation between traits [16, 17]. This method offers a foundation for understanding shared genetic architecture and complements causal inference approaches like LCV by offering insights into the degree of genetic overlap between traits.

Additionally, the LCV method is used to assess the potential causal genetic effects of two genetically correlated traits. LCV assumes a latent variable (L) that has a causal effect on each trait [11]. The genetic causality proportion (GCP) estimate indicates if the genetic correlation is due mainly to horizontal or vertical pleiotropy and is calculated by determining the correlations between latent variable L with trait A and trait B respectively [11]. The GCP estimate represents the proportion of a genetic correlation that could be described by potential causal effects. A GCP value of 0 indicates that the genetic correlation is mediated by horizontal pleiotropic effects, suggesting the absence of causal genetic effects. On the other hand, |GCP| = 1 denotes the presence of vertical pleiotropic effects as well as complete genetic causality. A |GCP| > 0.6 is regarded as a robust indicator of potential vertical pleiotropic effects [11]. To interpret LCV results, three items must be considered (i) the magnitude of genetic correlation, (ii) the magnitude of the GCP estimation, and (iii) the directionality of the GCP estimate. Furthermore, a negative GCP value between stroke and trait A indicates that trait A may have causal genetic effects on stroke, whereas a positive GCP value between stroke and trait A indicates that stroke may have causal genetic effects on trait A.

We implemented the phenome-wide analysis pipeline in the CTG-VL as described in previous studies [18-21]. We conducted LD score regression [16] and LCV analyses [11] to assess genetic correlations and potential putative causal links, respectively. Full details for utilising the publicly accessible phenome-wide analysis methodology can be found elsewhere [13]. The LCV was performed on all traits that showed a genetic correlation with stroke (false discovery rate, FDR<0.05). Similarly, we considered all GCP estimate values that were significant after multiple testing corrections (FDR<0.05).

### Mendelian randomization analysis

We performed bi-directional generalised Mendelian randomisation (GSMR) analyses to confirm the putative causal associations identified via LCV. Mendelian randomisation methods leverage independent genome-wide genetic variants (SNPs) as instrumental variables to identify causal relationships between exposure and outcome [22]. GSMR used the MR framework to perform summary-based Mendelian randomisation (SMR) analysis on each SNP instrument. This analysis considered the variability in sampling for each SNP and the linkage disequilibrium (LD) between SNPs. It then combined the causal estimates from all SNP instruments using generalised least squares method. GSMR estimates the causal relationship between causal traits (exposure) and stroke (outcome) and vice versa. Additionally, GSMR method employed HEIDI outlier method (*p-value* < 0.01) to exclude the putative pleiotropic outlier SNPs and unbiasedly, estimates the causal effect of exposures on outcomes [23].

### Gene-based association analysis

We used mBAT-combo, a set-based test, to identify potential causal genes associated with complex traits. The mBAT-combo method, which combines two statistical approaches, mBAT and fastBAT, is a powerful tool that can detect gene-trait associations when the gene contains SNPs on the same haplotype with opposing directions of effect [24]. The mBAT-combo uses GWAS summary statistics and linkage-disequilibrium reference to summarise genetic associations with a trait of interest at the gene level [24]. We performed mBAT-combo analysis for stroke and traits identified via LCV and GSMR to identify potential causal disease-associated genes. We adjusted the *P-value* using the Benjamini-Hochberg method (Adjusted *p-value <* 0.05) for all traits to identify shared genes between stroke and its traits with potential causal associations.

### Colocalization analysis

To determine whether the risk factors and stroke exhibited a shared a common genetic signal in a particular locus, we performed Bayesian colocalization analysis using the coloc.abf function in coloc R package (https://github.com/chr1swallace/coloc). In our analysis, dataset1 comprised the summary statistics of stroke, containing the cis-SNPs located within the genomic region surrounding the associated gene of stroke and risk factors included the summary statistics of risk factors, containing the cis-SNPs located within the genomic region surrounding the same gene which is shared with stroke. The colocalization analysis assumed a single causal variant in the locus for each trait and calculated posterior probabilities whereas, H0: no causal variant for either trait, H1: a causal variant for stroke only, H2: a causal variant for risk factor only, H3: distinct causal variants for stroke and risk factor, H4: a shared causal variant for both traits. We used posterior probability of H4 (P(H4) to assess evidence of colocalization, with P(H4)>0.8 considered strong evidence of highly colocalised [25].

### Functional enrichment analysis

We performed a functional enrichment analysis of the common genes shared between stroke and its risk factors using the ConsensusPathDB bioinformatics tool [26], for identification of the shared biological mechanisms or pathways between them. ConcensusPathDB is the largest repository for functional enrichment analysis of genes. We performed pathway enrichment analyses of the shared genes using database for pathways such as KEGG, BioCarta, Reactome, and Wikipathways. The enrichment analysis adjusted p-value generated from the hypergeometric test for multiple testing corrections. We considered the significant pathways which passed the adjusted *p-value* < 0.05.

## Results

### Identification of causal traits associated with the risk of stroke using latent causal variable method

We identified 262 traits genetically correlated with stroke (FDR<0.05; sTable 1). Among these genetically correlated traits, 14 could be explained via robust vertical pleiotropic effects with stroke (|GCP|> 0.60; FDR <0.05), including cardiovascular traits (atrial fibrillation, cardiac arrhythmias), metabolic trait (gamma-glutamyl transferase), blood cell trait (platelet crit), health conditions such as pulmonary embolism, blood clot in the lung, blood clot in the leg, deep venous thrombosis of lower extremities (Table 1, sTable 2).

**Table 1.** Traits with potential causal association with stroke. This table represents the phenotypes with FDR<.05 and robust genetic causal proportions (|GCP| > 0.60) with stroke.

| Trait | GCP | GCPse | GCPpval | rG | rGse | rGpval |
| --- | --- | --- | --- | --- | --- | --- |
| Self-reported atrial fibrillation | -0.97 | 0.03 | $8.97 \times 10^{-202}$ | 0.58 | 0.18 | $9.51 \times 10^{-4}$ |
| Self-reported pulmonary embolism | -0.79 | 0.15 | $1.37 \times 10^{-7}$ | 0.45 | 0.13 | $3.20 \times 10^{-4}$ |
| Blood clot in the lung diagnosed by a doctor | -0.75 | 0.17 | $2.46 \times 10^{-5}$ | 0.44 | 0.13 | $3.41 \times 10^{-4}$ |
| Cardiac arrhythmias | -0.72 | 0.19 | $2.99 \times 10^{-4}$ | 0.44 | 0.08 | $1.76 \times 10^{-7}$ |
| Blood clot in the leg diagnosed by a doctor | -0.74 | 0.18 | $5.25 \times 10^{-5}$ | 0.43 | 0.11 | $1.96 \times 10^{-4}$ |
| Medication: Gliclazide | -0.66 | 0.23 | $3.75 \times 10^{-3}$ | 0.42 | 0.09 | $2.33 \times 10^{-5}$ |
| Self-reported deep venous thrombosis | -0.74 | 0.18 | $4.23 \times 10^{-5}$ | 0.40 | 0.11 | $2.60 \times 10^{-4}$ |
| Atrial fibrillation and flutter (ICD10) | -0.80 | 0.14 | $2.10 \times 10^{-8}$ | 0.40 | 0.08 | $1.91 \times 10^{-6}$ |
| Medication: Insulin | -0.84 | 0.12 | $1.43 \times 10^{-11}$ | 0.38 | 0.14 | $6.43 \times 10^{-3}$ |
| Medication: Pioglitazone | -0.74 | 0.17 | $1.72 \times 10^{-5}$ | 0.33 | 0.12 | $8.45 \times 10^{-3}$ |
| Weight change during worst episode of depression: Gained weight | -0.64 | 0.23 | $4.03 \times 10^{-3}$ | 0.32 | 0.12 | $1.01 \times 10^{-2}$ |
| Deep venous thrombosis of lower extremities and pulmonary embolism | -0.78 | 0.14 | $1.28 \times 10^{-8}$ | 0.24 | 0.08 | $6.00 \times 10^{-3}$ |
| Gamma glutamyltransferase (female) | -0.69 | 0.15 | $4.13 \times 10^{-6}$ | 0.16 | 0.04 | $3.23 \times 10^{-4}$ |
| Platelet crit | -0.67 | 0.20 | $7.61 \times 10^{-4}$ | 0.14 | 0.04 | $1.32 \times 10^{-3}$ |
| Gamma glutamyltransferase (male) | -0.64 | 0.19 | $7.61 \times 10^{-4}$ | 0.14 | 0.05 | $6.60 \times 10^{-3}$ |

This table represents the traits with significant false discovery rate ( $FDR < .05$ ); GCP: genetic causal proportion; GCP se: genetic causal proportion standard deviation; GCP pval: genetic causal proportion unadjusted p-value; rG: genetic correlation; rG se: genetic correlation standard deviation; rG pval: genetic correlation unadjusted p-value.

### Exploring the relationships between causal traits and stroke using generalized summary-based Mendelian randomization

To further support the causal relationships identified by LCV method, we performed generalized summary-based Mendelian randomization (GSMR) analysis considering putative risk traits identified via the LCV as the exposure and stroke as an outcome. We observed consistent findings by both methods, demonstrating a significant causal effect of risk traits on stroke. Specifically, GSMR suggested potential causal genetic effects of cardiovascular traits (atrial fibrillation, cardiac arrhythmias), metabolic traits (gamma-glutamyl transferase), blood cell traits (platelet crit), and health conditions (pulmonary embolism, blood clot in the lung, blood clot in the leg, deep venous thrombosis of lower extremities), on a higher risk of stroke (Table 2). However, no evidence of a reverse causal relationship was observed (Table 3).

**Table 2.** Traits with potential causal association with stroke using generalized Mendelian randomization analysis.

| Exposure | Outcome | Effect | SE | *P* | *P* HEIDI | SNPs |
| --- | --- | --- | --- | --- | --- | --- |
| Self-reported atrial fibrillation | Stroke | 13.80 | 1.37 | $1.2 \times 10^{-23}$ | 0.28 | 13 |
| Self-reported pulmonary embolism | | 20.06 | 2.75 | $3.2 \times 10^{-13}$ | 0.31 | 4 |
| Blood clot in the lung diagnosed by a doctor | | 17.20 | 2.69 | $1.6 \times 10^{-10}$ | 0.12 | 3 |
| Cardiac arrhythmias | | 7.00 | 0.65 | $1.4 \times 10^{-26}$ | 0.06 | 24 |
| Blood clot in the leg diagnosed by a doctor | | 4.72 | 1.13 | $2.9 \times 10^{-5}$ | 0.1 | 9 |
| Medication: Gliclazide |  | 5.23 | 2.22 | 0.01 | 0.08 | 7 |
| Self-reported deep venous thrombosis | | 4.75 | 1.14 | $3.2 \times 10^{-5}$ | 0.09 | 9 |
| Atrial fibrillation and flutter (ICD10) | | 7.40 | 0.66 | $6.1 \times 10^{-29}$ | 0.12 | 29 |
| Medication: Insulin |  | 1.89 | 0.89 | 0.02 | 0.2 | 10 |
| Deep venous thrombosis of lower extremities and pulmonary embolism | | 15.21 | 2.18 | $3.49 \times 10^{-12}$ | 0.26 | 5 |
| Gamma glutamyltransferase | | 0.07 | 0.01 | $8.23 \times 10^{-7}$ | 0.05 | 654 |
| Platelet crit | | 1.72 | 0.37 | $4.71 \times 10^{-6}$ | 0.06 | 616 |

This table represents the traits with significant false discovery rate (FDR < .05); SE: standard error; *P*: *p*-value; HEIDI test *P*: heterogeneity in dependent instruments test. SNPs#: number of single nucleotide polymorphism.

**Table 3.**
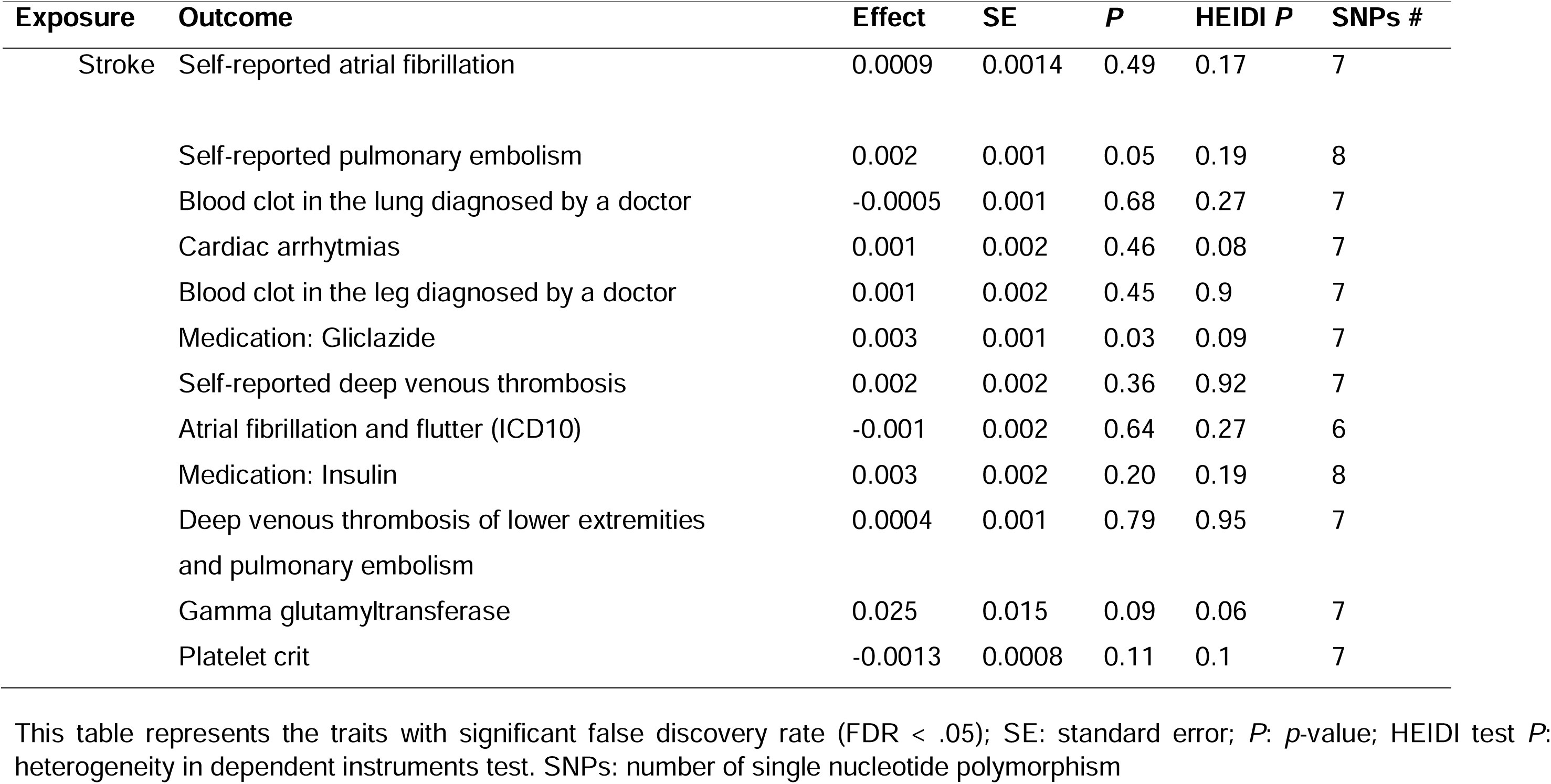
Reverse generalized Mendelian randomization analysis between exposure stroke and causal traits as outcome.

### Gene-based association analysis of causal traits and stroke

Using mBAT-combo, we identified 72 genes with stroke, 70 genes associated with atrial fibrillation, 267 genes with atrial fibrillation and flutter, 115 genes with blood clot in the leg, 69 genes with blood clot in lung, 192 genes with cardiac arrhymias, 77 genes with DVT, 103 genes with DVT of lower extremities and pulmonary embolism, 44 genes with pulmonary embolism, 8,568 genes with platelet crit, and 6,443 genes with gamma-glutamyl transferase, respectively (*adjusted p-value <0.05)* (sTable 3 to sTable 12). Several overlapping genes between causal traits and stroke were identified (sTable 13). Notably, *ZFHX3, OBFC1,* and *SH3PXD2A* were shared between stroke and atrial fibrillation or cardiac arrhythmias. Additionally, *FGA, FGG, ILF3, FGB, ILF3-AS,* and *SH2B3* were associated with blood clots in the leg, deep venous thrombosis in the lower extremities, pulmonary embolism, and stroke (Figure 3.2 and sTable 13).

### Colocalization analysis of shared genes in stroke and risk factors

We identified several colocalised genes (PPH4 > 80%) between stroke and its associated risk factors (sTable 13). The high PPH4 values suggest that these loci are likely to influence both stroke and its associated risk factors through shared genetic mechanisms. Colocalised genes included *ZFHX3* (PPH4>□95%), which was colocalised with atrial fibrillation and stroke, while *OBFC1* (PP.H4 > 95%) and *ZFHX3* (PP.H4 >91%) colocalised with cardiac arrhythmias and stroke. We also identified several other genes, *FGA*, *FGB,* and *FGG*, which were colocalized with deep venous thrombosis, pulmonary embolism, blood clot in the lung, and blood clot in the leg (PP.H4 >98%; sTable 13). We observed *CDK6, FAM133B, FAM133DP, ACAD10, ALDH2, ERP29, QTRT1* and some other genes which are colocalised with platelet crit and stroke with PP.H4 > 98% (sTable 13).

### Functional enrichment analysis

To understand the biological functions and molecular pathways that underlie shared genetic overlap between causal traits and stroke, we performed a functional enrichment analysis of those shared candidate genes. We identified “intrinsic and extrinsic prothrombin activation pathway”, and “fibrinolysis pathway” to be associated with atrial fibrillation, cardiac arrhythmias, and stroke (Figure 3). In addition, we observed “complement and coagulation cascade”, “platelet activation”, “fibrinolysis”, “neutrophil extracellular trap formation” associated with blood clot in lung, blood clot in leg, deep venous thrombosis, pulmonary embolism, and stroke. Furthermore, “Hepatocellular carcinoma”, “COVID-19 disease”, “Oxytocin” and among other pathways were shared between platelet crit and stroke, (sTable 14 to sTable 22).

**Figurer 2.**
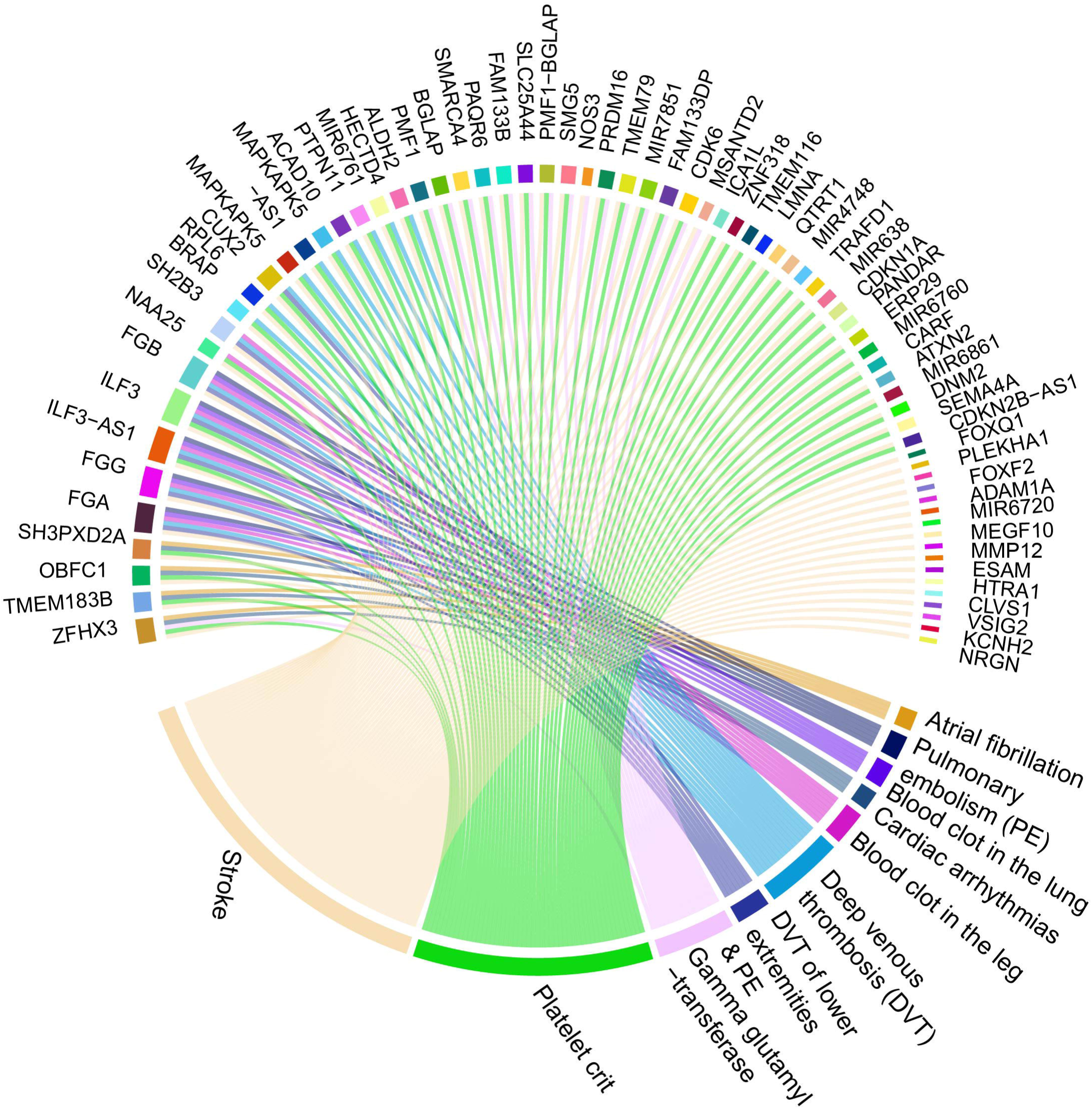
The chord diagram represents the genetic architecture linking stroke to its causal traits. Right side represents causal traits and stroke, and left side represents shared genes between causal traits and stroke. The These findings provide a basis for future functional studies to validate the roles of these shared genes in stroke pathogenesis.

**Figure 3.**
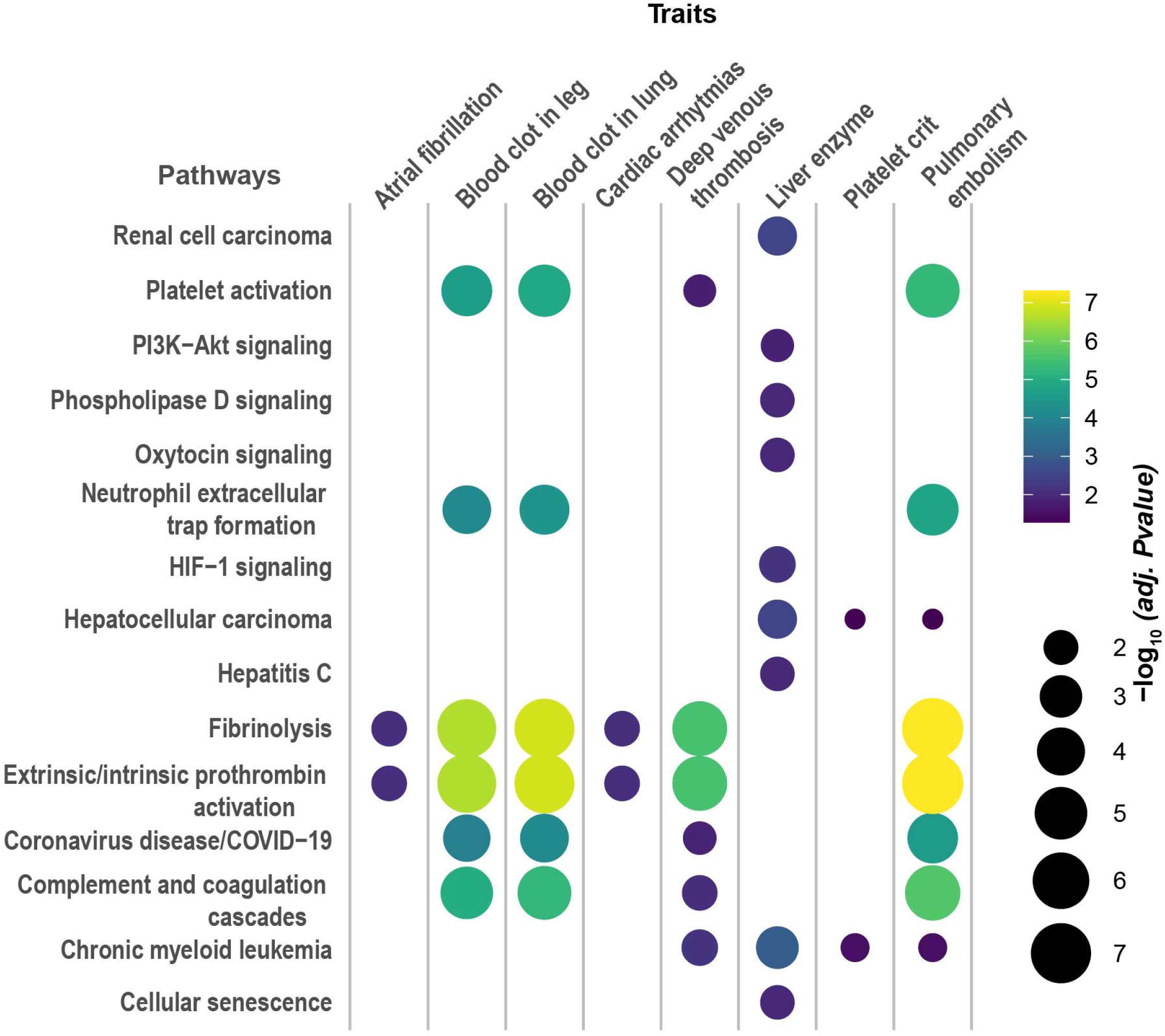
The bubble plot represents the common biological pathways shared between complex traits and stroke. The pathways are listed on the left, and the traits associated with stroke are displayed along the top. Each bubble represents a significant association between a pathway and a trait, with the size of the bubble corresponding to the statistical significance of the association (measured as -log10 of the adjusted p-value) and the color scale indicating the strength of the p-value (yellow = more significant, purple = less significant).

## Discussion

Our findings support some of the potential causal architectures hypothesized by observational studies and provide molecular factors underlying stroke and its causal traits, specifically cardiovascular, metabolic, and blood clot-related traits. Risk genes and pathways highlight the role of coagulation and complement systems, which underlie the shared biology between stroke and risk factors. Emerging studies have suggested that complement systems may serve as the coordinator of the cascade of molecular events in brain ischemia [27, 28], suggesting that these pathways may be potential targets for future intervention and treatment in managing stroke risk.

The findings of this study suggest a potential causal association of factors that may contribute to the risk of stroke, specifically associated with cardiovascular function, coagulation, and metabolic pathways. Although observational studies have demonstrated an association between some of these factors and the risk of stroke, these associations are often confounded by reverse causation, leaving the underlying genetic mechanisms and molecular factors incompletely understood. This study provides novel genetic causal architectures linking stroke and its risk factors. Metabolic dysregulation has emerged as a significant contributor to stroke susceptibility, with hepatic gamma-glutamyl transferase (GGT) playing a crucial role in linking metabolic dysfunction to vascular health. GGT is a hepatic biomarker linked to liver dysfunction, physiology, and metabolism [29]. Genetic variants associated with GGT are correlated with increased risk of cardiovascular diseases and lipid and glucose metabolism [29]. Enhanced GGT may increase oxidative stress and contribute to endothelial dysfunction and atherogenesis, ultimately contributing to the elevated stroke risk. This is consistent with molecular pathways enriched in various pathways implicated in oxidative damage, blood coagulation, and thrombosis processes, all of which ultimately may lead to neuronal deaths [30, 31]. Further, gene-based association analyses revealed shared genetic architecture between stroke and causal traits.

Our findings suggest overlapping risk genes between stroke and causal traits. For instance, *FGA, FGB,* and *FGG* were found overlapping among stroke and deep venous thrombosis, blood clots in the lungs and legs, and pulmonary embolisms. Each of these genes encodes fibrinogen protein, such as *FGA* encodes fibrinogen Aα chain protein*, FGB* encodes fibrinogen Bβ chain protein, and *FGG* encodes fibrinogen γ chain protein [25]. Fibrinogen has a significant role in the coagulation process, particularly in response to any injury where fibrinogen is transformed into fibrin, the main component of blood clots. *FGA*, *FGB,* and *FGG* were proposed as associated with ischemic stroke [32, 33]. Likewise, we observed *ZFHX3* overlapped between stroke and atrial fibrillation, atrial fibrillation flutter, gamma glutamyl-transferase, platelet crit, and cardiac arrhythmias. *ZFHX3* is a zinc-finger homeodomain transcription factor associated with numerous biological processes, including cell differentiation and tumorigenesis. A previous study reported that a sequence variant in *ZFHX3* on 16q22 is associated with the pathogenesis of atrial fibrillation and ischemic stroke [34]. Furthermore, *OBFC1 and SH3PXD2A* overlapped atrial fibrillation and cardiac arrhythmias with stroke. These genes have been proposed as the putative causal genes in our previous study [35].

One limitation of our study is that we have included medication-based GWAS datasets for some traits as a proxy to investigate disease risk. While medication GWAS as proxy can provide critical insights into the genetic mechanism of the disease but it may reflect aspects of treatment response rather than the disease itself. Additionally, the more substantial statistical power of medication-use GWAS (pioglitazone, insulin, and gliclazide) compared to disease-based GWAS (e.g., type 2 diabetes) may explain why we did not observe a significant association between non-medication-based GWAS for type 2 diabetes and stroke risk. Future studies employing the most robust direct disease GWAS may help disentangle the genetic contributions of disease susceptibility from treatment effects. Additionally, we acknowledge that the findings of this study have focused on European-based GWAS datasets. Therefore, future studies will be required to validate these findings in diverse populations and investigate the potential for translating these findings into clinical applications.

## Conclusion

In summary, our study provides evidence of potential causal genetic effects and shared genetic architecture between stroke and risk factors, including cardiovascular, metabolic, and blood clot-related traits. These findings suggest potential molecular mechanisms that link genetic predispositions to stroke risk, which in turn could be leveraged to improve the development of future studies and preventative strategies.

## Data Availability Statement

All data here analyzed is publicly available on MEGASTROKE consortium and the UK biobank summary data is available on CTG-VL.

## Acknowledgments

We thank the MEGASTROKE project for making stroke GWAS summary statistics data publicly available. The author list of MEGASTROKE is available in Supplementary Table 23. T.I. is supported by a Research Training scholarship from The University of Queensland, Australia.

## Author contributions

TI, JZ, MAM contributed to the study conception, design critical reading and editing. TI performed bioinformatics analysis, prepared the figures and wrote the manuscript. LMGM, MER, GCP, AK interpretation and critical reading.

## Supplementary Tables

Supplementary Tables 1-23

## References

1. Organization, W.H., World health statistics 2010. 2010: World Health Organization.

2. Kuriakose, D. and Z. Xiao, Pathophysiology and Treatment of Stroke: Present Status and Future Perspectives. Int J Mol Sci, 2020. 21(20).

3. Dichgans, M., Genetics of ischaemic stroke. Lancet Neurology, 2007. 6(2): p. 149–161.

4. Bevan, S., et al., Genetic Heritability of Ischemic Stroke and the Contribution of Previously Reported Candidate Gene and Genomewide Associations. Stroke, 2012. 43(12): p. 3161-+.

5. Gretarsdottir, S., et al., Risk variants for atrial fibrillation on chromosome 4q25 associate with ischemic stroke. Ann Neurol, 2008. 64(4): p. 402–9.

6. Liu, J.F., et al., Causal Impact of Type 2 Diabetes Mellitus on Cerebral Small Vessel Disease: A Mendelian Randomization Analysis. Stroke, 2018. 49(6): p. 1325–1331.

7. Georgakis, M.K. and D. Gill, Mendelian Randomization Studies in Stroke Exploration of Risk Factors and Drug Targets With Human Genetic Data. Stroke, 2021. 52(9): p. 2992–3003.

8. García-Marín, L.M., et al., Phenome-wide analysis highlights putative causal relationships between self-reported migraine and other complex traits. Journal of Headache and Pain, 2021. 22(1).

9. Siewert, K.M., et al., Cross-trait analyses with migraine reveal widespread pleiotropy and suggest a vascular component to migraine headache. International Journal of Epidemiology, 2020. 49(3): p. 1022–1031.

10. van Rheenen, W., et al., Genetic correlations of polygenic disease traits: from theory to practice. Nature Reviews Genetics, 2019. 20(10): p. 567–581.

11. O’Connor, L.J. and A.L. Price, Distinguishing genetic correlation from causation across 52 diseases and complex traits. Nature Genetics, 2018. 50(12): p. 1728-+.

12. Koellinger, P.D. and R. de Vlaming, Mendelian randomization: the challenge of unobserved environmental confounds. International Journal of Epidemiology, 2019. 48(3): p. 665–671.

13. Haworth, S., et al., Assessment and visualization of phenome-wide causal relationships using genetic data: an application to dental caries and periodontitis. European Journal of Human Genetics, 2021. 29(2): p. 300–308.

14. Malik, R., et al., Multiancestry genome-wide association study of 520,000 subjects identifies 32 loci associated with stroke and stroke subtypes. Nature Genetics, 2018. 50(4): p. 524-+.

15. Cuellar-Partida, G., et al., Complex-Traits Genetics Virtual Lab: A community-driven web platform for post-GWAS analyses. BioRxiv, 2019: p. 518027.

16. Bulik-Sullivan, B.K., et al., LD Score regression distinguishes confounding from polygenicity in genome-wide association studies. Nature Genetics, 2015. 47(3): p. 291-+.

17. Bulik-Sullivan, B., et al., An atlas of genetic correlations across human diseases and traits. Nat Genet, 2015. 47(11): p. 1236–41.

18. García-Marín, L.M., et al., Inference of causal relationships between sleep-related traits and 1,527 phenotypes using genetic data. Sleep, 2021. 44(1).

19. García-Marín, L.M., et al., Phenome-wide screening of GWAS data reveals the complex causal architecture of obesity. Human Genetics, 2021. 140(8): p. 1253–1265.

20. García-Marín, L.M., et al., Large-scale genetic investigation reveals genetic liability to multiple complex traits influencing a higher risk of ADHD. Scientific Reports, 2021. 11(1).

21. García-Marín, L.M., et al., Shared molecular genetic factors influence subcortical brain morphometry and Parkinson’s disease risk. Npj Parkinsons Disease, 2023. 9(1).

22. Davey Smith, G. and S. Ebrahim, ‘Mendelian randomization’: can genetic epidemiology contribute to understanding environmental determinants of disease? International journal of epidemiology, 2003. 32(1): p. 1–22.

23. Zhu, Z.H., et al., Causal associations between risk factors and common diseases inferred from GWAS summary data. Nature Communications, 2018. 9.

24. Li, A., et al., mBAT-combo: A more powerful test to detect gene-trait associations from GWAS data. American Journal of Human Genetics, 2023. 110(1): p. 30-+.

25. Lin, J.F., J.W. Zhou, and Y. Xu, Potential drug targets for multiple sclerosis identified through Mendelian randomization analysis. Brain, 2023. 146(8): p. 3364–3372.

26. Kamburov, A., U. Stelzl, H. Lehrach, and R. Herwig, The ConsensusPathDB interaction database: 2013 update. Nucleic Acids Research, 2013. 41(D1): p. D793–D800.

27. Fumagalli, S. and M.G. De Simoni, Lectin Complement Pathway and Its Bloody Interactions in Brain Ischemia. Stroke, 2016. 47(12): p. 3067–3073.

28. Kenawy, H.I., I. Boral, and A. Bevington, Complement-coagulation cross-talk: a potential mediator of the physiological activation of complement by low pH. Frontiers in Immunology, 2015. 6.

29. Pazoki, R., et al., Genetic analysis in European ancestry individuals identifies 517 loci associated with liver enzymes. Nature Communications, 2021. 12(1).

30. Grover, S.P. and N. Madman, Intrinsic Pathway of Coagulation and Thrombosis: Insights From Animal Models. Arteriosclerosis Thrombosis and Vascular Biology, 2019. 39(3): p. 331–338.

31. Doyle, K.P., R.P. Simon, and M.P. Stenzel-Poore, Mechanisms of ischemic brain damage. Neuropharmacology, 2008. 55(3): p. 310–318.

32. Jood, K., et al., Fibrinogen gene variation and ischemic stroke. Journal of Thrombosis and Haemostasis, 2008. 6(6): p. 897–904.

33. Ken-Dror, G., et al., Mendelian randomization assessing causal relationship between fibrinogen levels and ischemic stroke. Journal of Stroke & Cerebrovascular Diseases, 2025. 34(2).

34. Gudbjartsson, D.F., et al., A sequence variant in ZFHX3 on 16q22 associates with atrial fibrillation and ischemic stroke. Nature Genetics, 2009. 41(8): p. 876–878.

35. Islam, T., M.R. Rahman, A. Khan, and M.A. Moni, Integration of Mendelian randomisation and systems biology models to identify novel blood-based biomarkers for stroke. Journal of Biomedical Informatics, 2023. 141.

